# Top 100 cited systematic reviews and meta-analyses in the major journals of oral and maxillofacial surgery : a bibliometric analysis

**DOI:** 10.1101/2020.12.02.20242503

**Authors:** Essam Ahmed Almoraissi, Endi Lanza Galvão, Saulo Nikolaos Christidis, Gabriel Moreira Falci

## Abstract

The aim of this bibliometric research was to identify and analyze the top 100 cited systematic reviews in the field of oral and maxillofacial surgery. Using the Web of Science-database without restrictions on publication year or language, a bibliometric analysis was performed for the five major journals of oral and maxillofacial surgery: International Journal of Oral and Maxillofacial Surgery, Journal of Oral and Maxillofacial Surgery, Journal of Cranio-maxillofacial Surgery, British Journal of Oral & Maxillofacial Surgery and Oral Surgery Oral medicine Oral pathology Oral radiology. The most top-cited systematic review was published in 2015 with a total of 200 citations on survival and success rates of dental implants, consistent with the finding that “pre- and peri-implant surgery and dental implantology”, and “craniomaxillofacial deformities and cosmetic surgery” were the most frequently-cited topics (22% each). The International Journal of Oral and Maxillofacial Surgery and Journal of Oral and Maxillofacial Surgery displayed have got most citations in total and in average per publication. The outcome of this article can be used as a source of information not just for researchers but also for clinicians and students, and of which areas have a large impact on the field of oral and maxillofacial surgery but cannot reflect the quality of the included systematic reviews.

## INTRODUCTION

Decision-making regarding all clinical decisions including choice of treatment is nowadays nationally and internationally based on evidence-based medicine in all fields of medicine such as dentistry and oral and maxillofacial surgery (OMFS) ^1,2^. Randomized controlled trials (RCTs) have been considered the best and strongest study design to answer a specific clinical question, consequently being a guide to the decision making, described as an evidence-based approach. However, in most of the RCTs, a huge sample size is warranted to reach an outcome based on reliable statistics. Performing a RCT with a big sample size is a challenge for any researcher. To solve this problem, a study design called systematic review with meta-analysis was proposed. This type of research allows the researchers to agglutinate data extracted from many RCTs, in only one statistical analysis. Thus, this kind of statistical analysis can provide a more robust and stronger outcome/result about one specific clinical question. Besides that, there are tools to evaluate the quality of RCTs, in order to be able to do recommendations about this specific clinical issue. Therefore, presently systematic reviews and meta-analysis are graded with the highest quality level study design ^3^.

Another type of research, used to show which impact publications have, is called “bibliometrics”. The first citation about bibliometrics was done by Pritchard in 1969 ^4^. There are some publications on bibliometric analysis in the field of OMSF, but specified as facial trauma ^5^, oral cancer ^6^, and maxillofacial surgery ^7^. The citation analysis is a type of bibliometrics which quantifies how many times a publication has been cited after its publication. This information can be efficient to use in order to evaluate which impact a publication has in a specific field, and therefore how important this publication is in the specific field. Thus, the more cited the publication is, the greater ability it has to influence clinical decisions ^8^. Furthermore, this analysis is an important tool to help the clinicians to identify the best publications in their field of action and to guide them in their decision making based on these publications.

After the millennial shift there was a huge increase in the number of published systematic reviews in medicine and dentistry. Specifically, in the field of OMFS the increase of published systematic reviews was observed after 2010. To our knowledge there are no studies aiming to investigate the most cited systematic reviews in the field of OMFS. Thus, after 10 years of worldwide research it is imperative to know how the systematic reviews are influencing the decision making in the field of OMFS, and to rank the most cited systematic reviews in order to provide a guidance to the clinicians acquisition of knowledge. The aim of this bibliometric research was therefore to identify and analyze the top 100 cited systematic reviews in the field of OMFS.

## METHODS

The authors performed a bibliometric analysis of the top 100 most highly cited systematic reviews in the five major journals of oral and maxillofacial surgery namely: International Journal of Oral and Maxillofacial Surgery (IJOMS), Journal of Oral and Maxillofacial Surgery (JOMS), Journal of Cranio-maxillofacial Surgery (JCMS), British Journal of Oral & Maxillofacial Surgery (BJOMS) and Oral Surgery Oral medicine Oral pathology Oral radiology (TRIPLEO). A search was performed on 29^th^ October 2020, using the Clarivate Analytics’ Web of Science database and there was no restriction on the publication year or the language of the manuscripts. The search strategy was “systematic review” OR “systematic reviews” OR “meta-analysis” OR “meta-analyses”. The results were organized in descending order of citation count.

Every systematic review was stratified into the following categories: dentoalveolar surgery, pre-implant surgery and dental implantology, traumatology, craniomaxillofacial deformities and cosmetic surgery, osteonecrosis of the jaws, pathology, reconstructive surgery, temporomandibular joint (TMJ), basic science research, and emerging technologies

The most cited articles were analyzed regarding the following information: number of citations, publication year, journals, authors, number of authors, methodological design (systematic review or systematic review with meta-analysis), article topic, contributing institution and country. The country of origin and contributing institution of the article was defined by the address provided for the corresponding author. When the paper presented the same citation number the youngest was best ranked.

Number of articles and citations per article were graphed using the Statistical Package for the Social Sciences software (SPSS version 22.0).

## RESULTS

The initial search identified 771 articles. The 100 top-cited systematic reviews on the OMFS field are listed by rank order based on the number of citations in **Table 1**. From a total of 100 systematic reviews only 37 presented meta-analyses. The number of authors ranged between one and 16 (mean 4.12 ± 2.31).

**Table 1.**
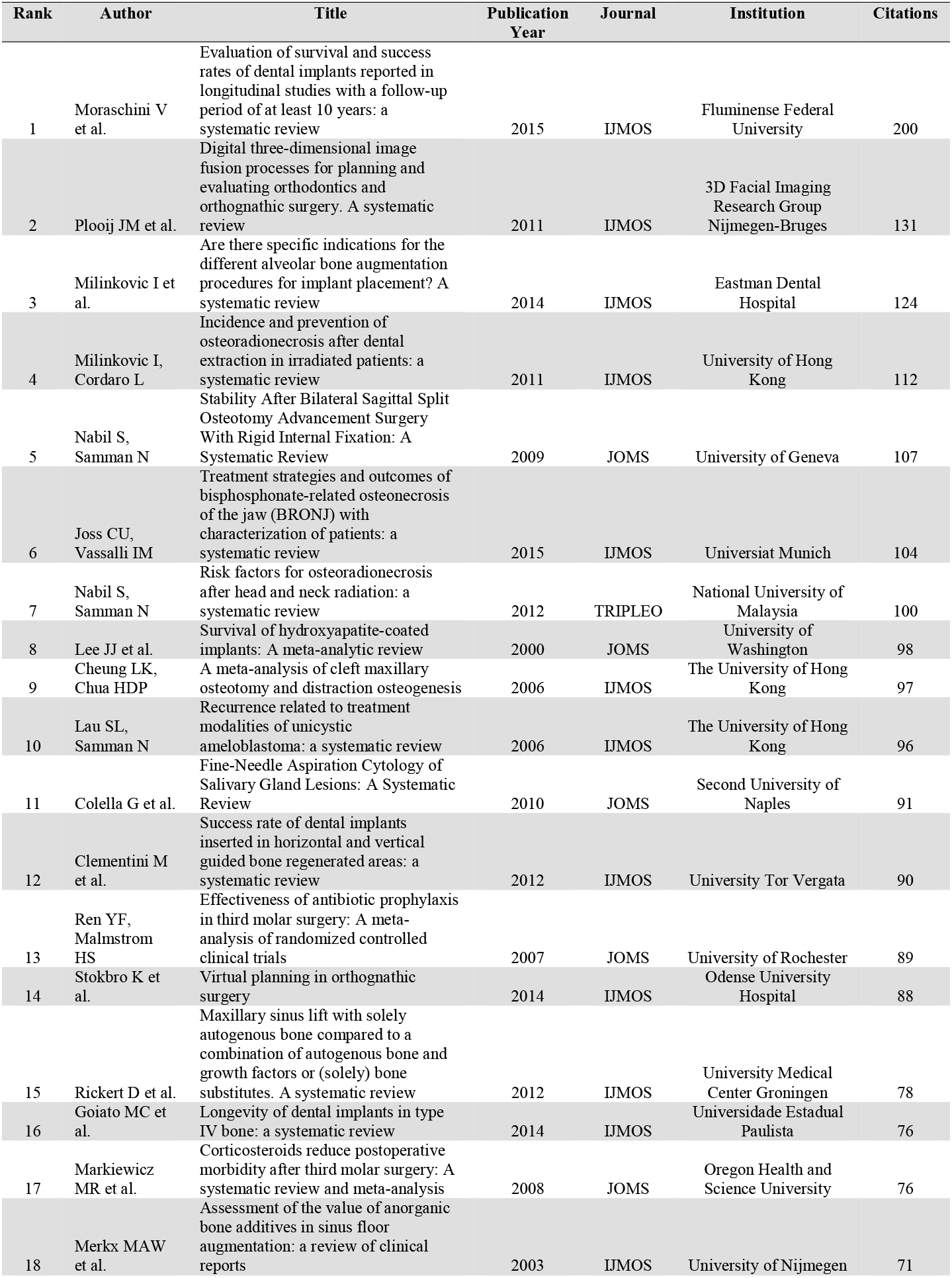
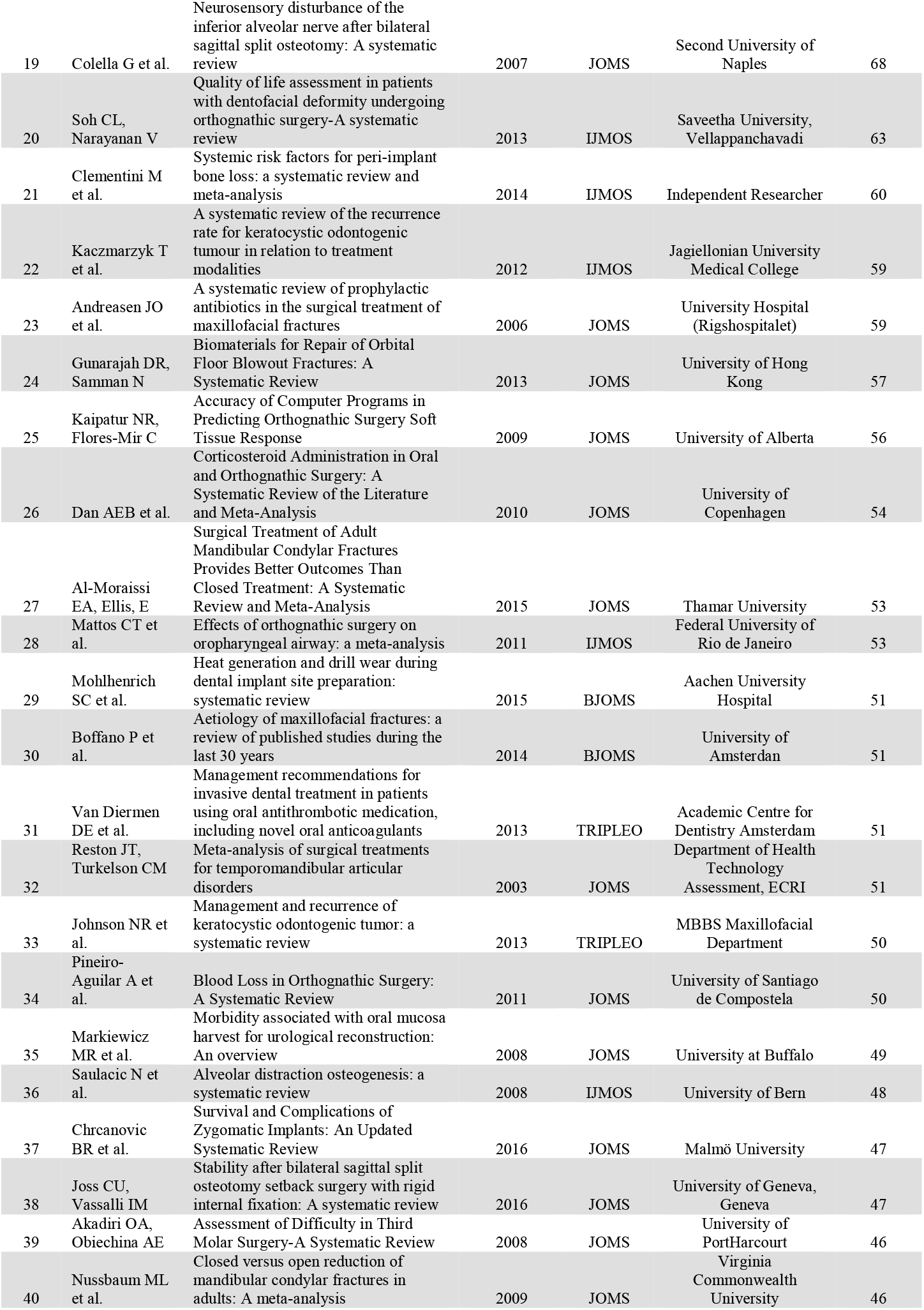
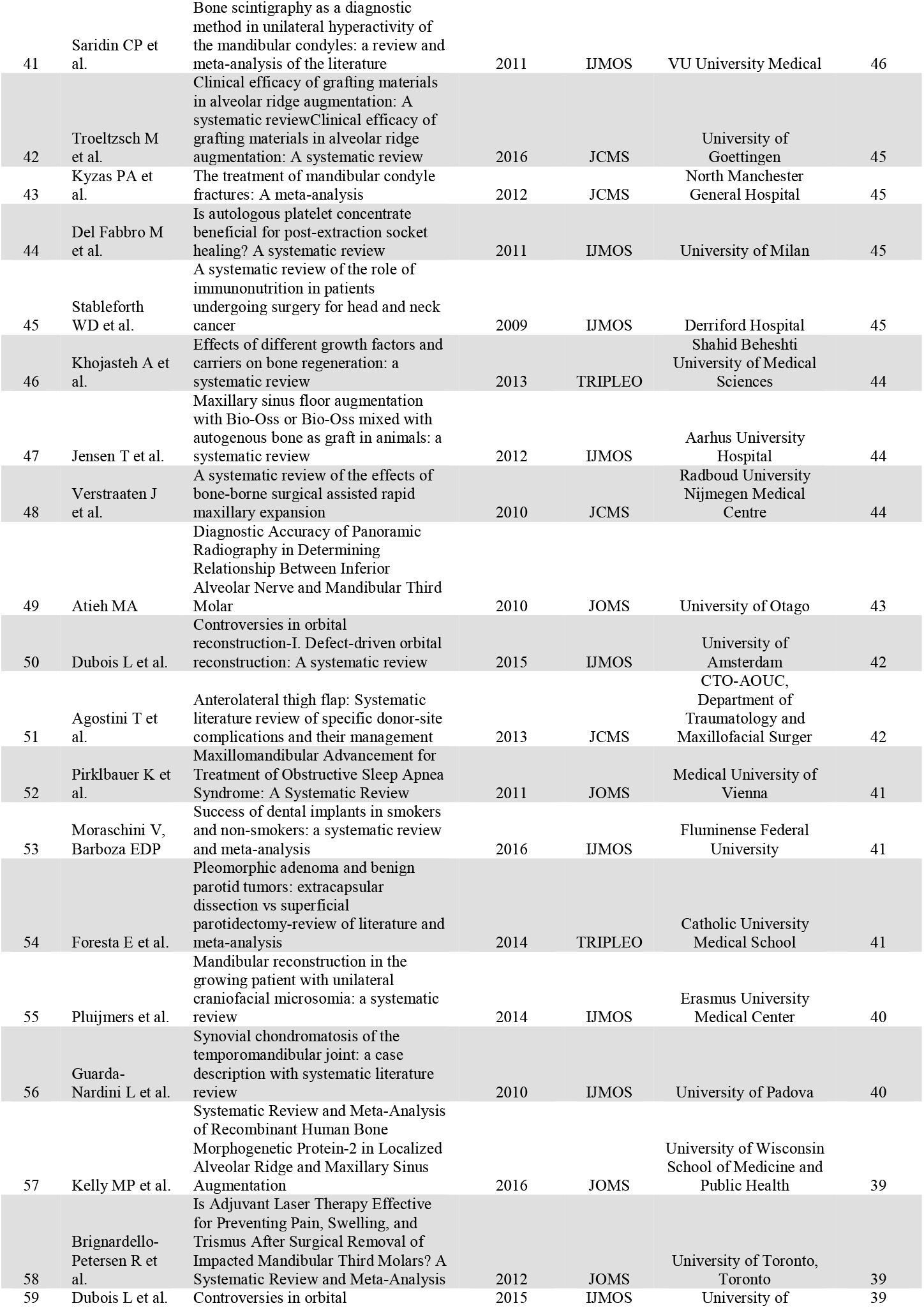
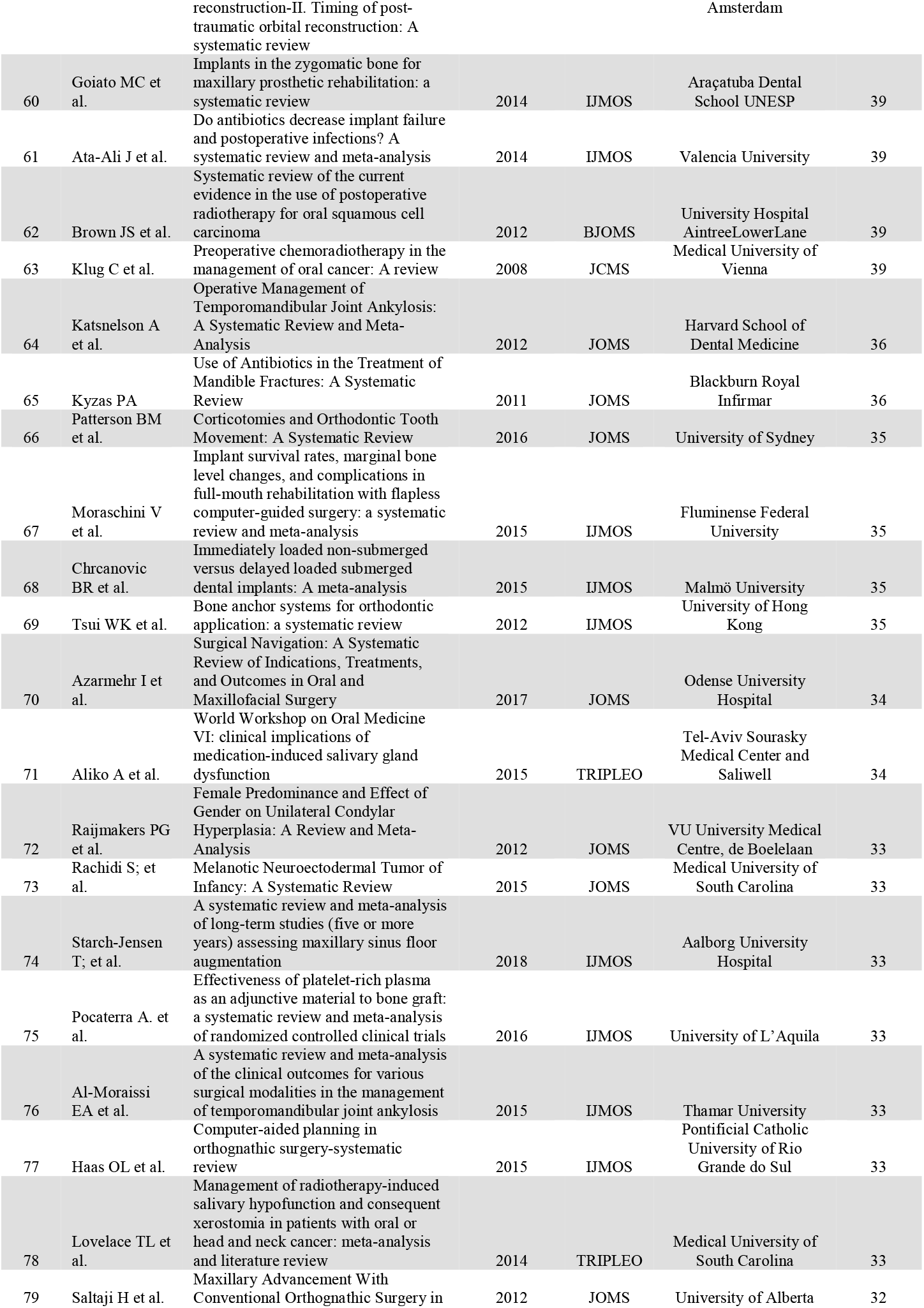
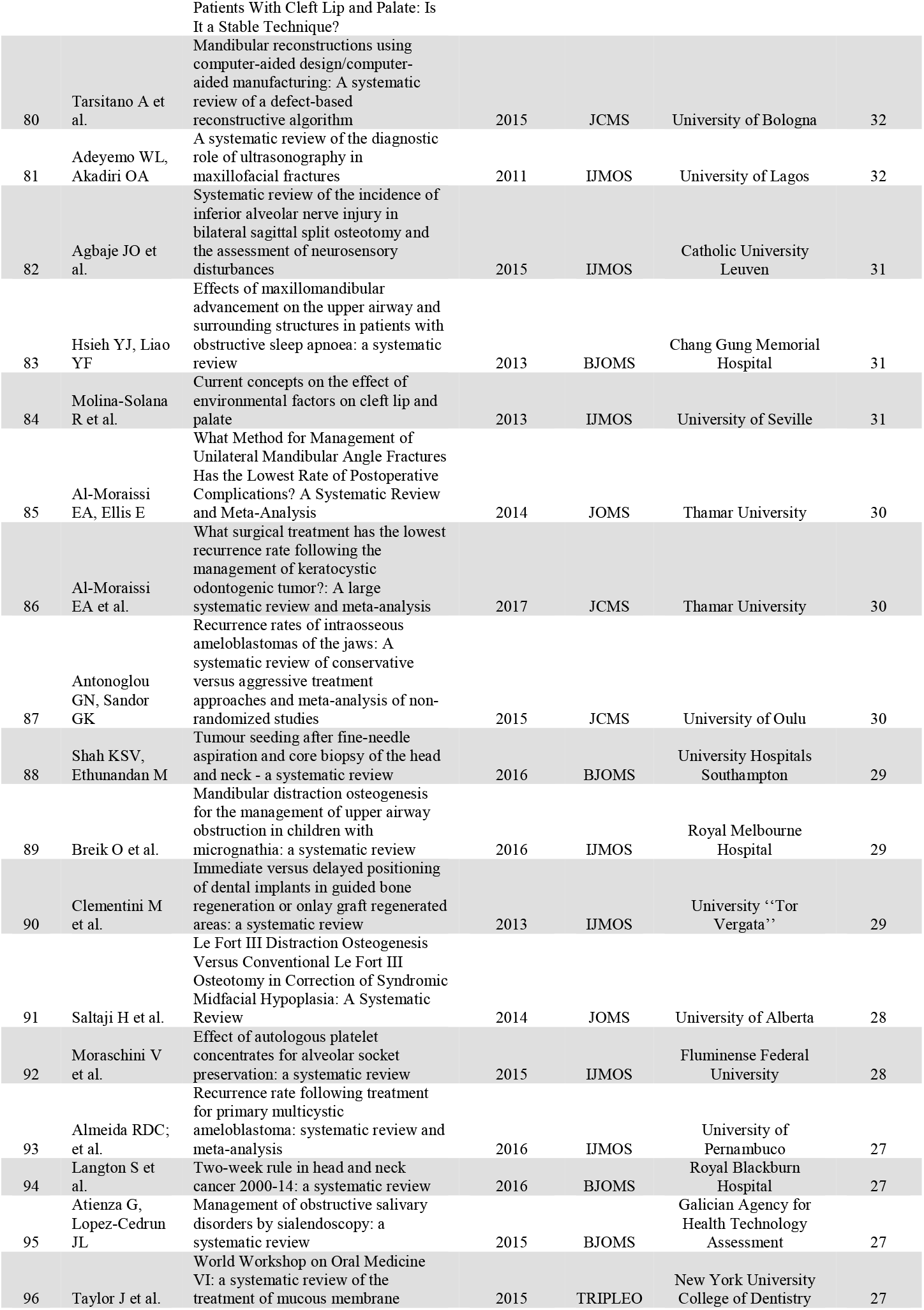
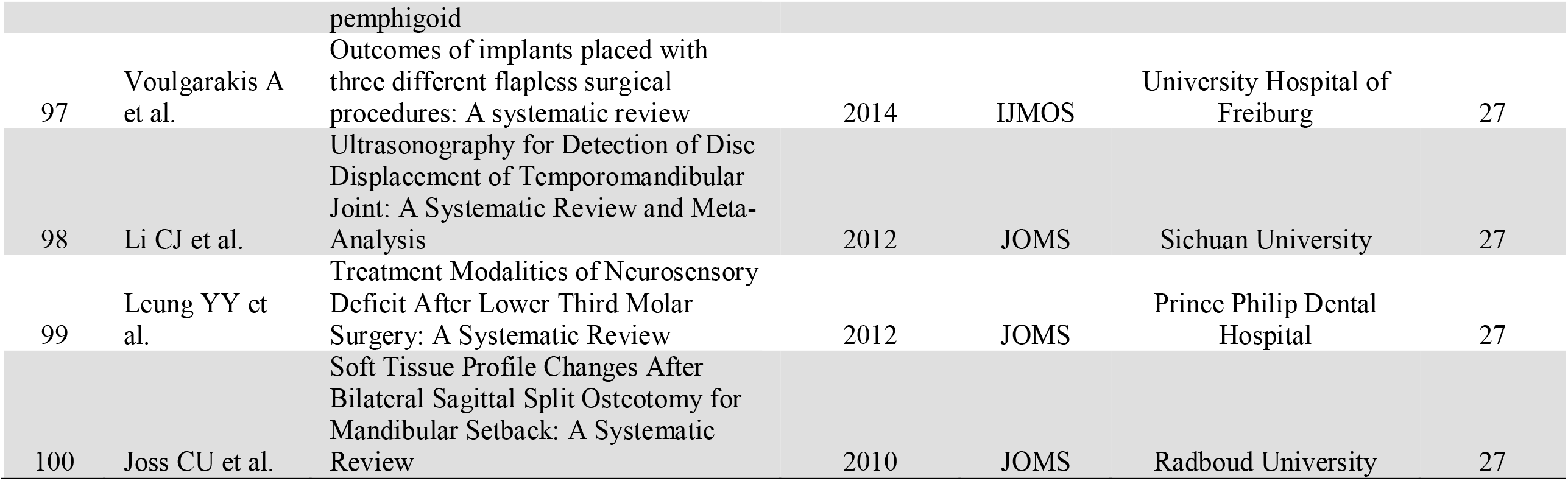
The top 100 cited systematic review and meta-analyses in oral and maxillofacial surgery.

These articles have been cited a combined total of 5107 times. The most top-cited article was published in 2015 with a total of 200 citations ^9^. Based on the distribution of the 100 articles over the years and their citations per publication, the years 2000 followed by 2006 and 2007 were the most productive years (**Figure 1**). The earliest systematic review included in this bibliometric analysis was published in 2000 by Lee et al.^10^, in JOMS and has been cited 98 times, while the most recently was published in 2018 by Starch-Jensen et al. ^11^in the IJOMS and has been cited 33 times. **Figure 2** illustrates the distribution of the 100 articles over the years.

**Figure 1.**
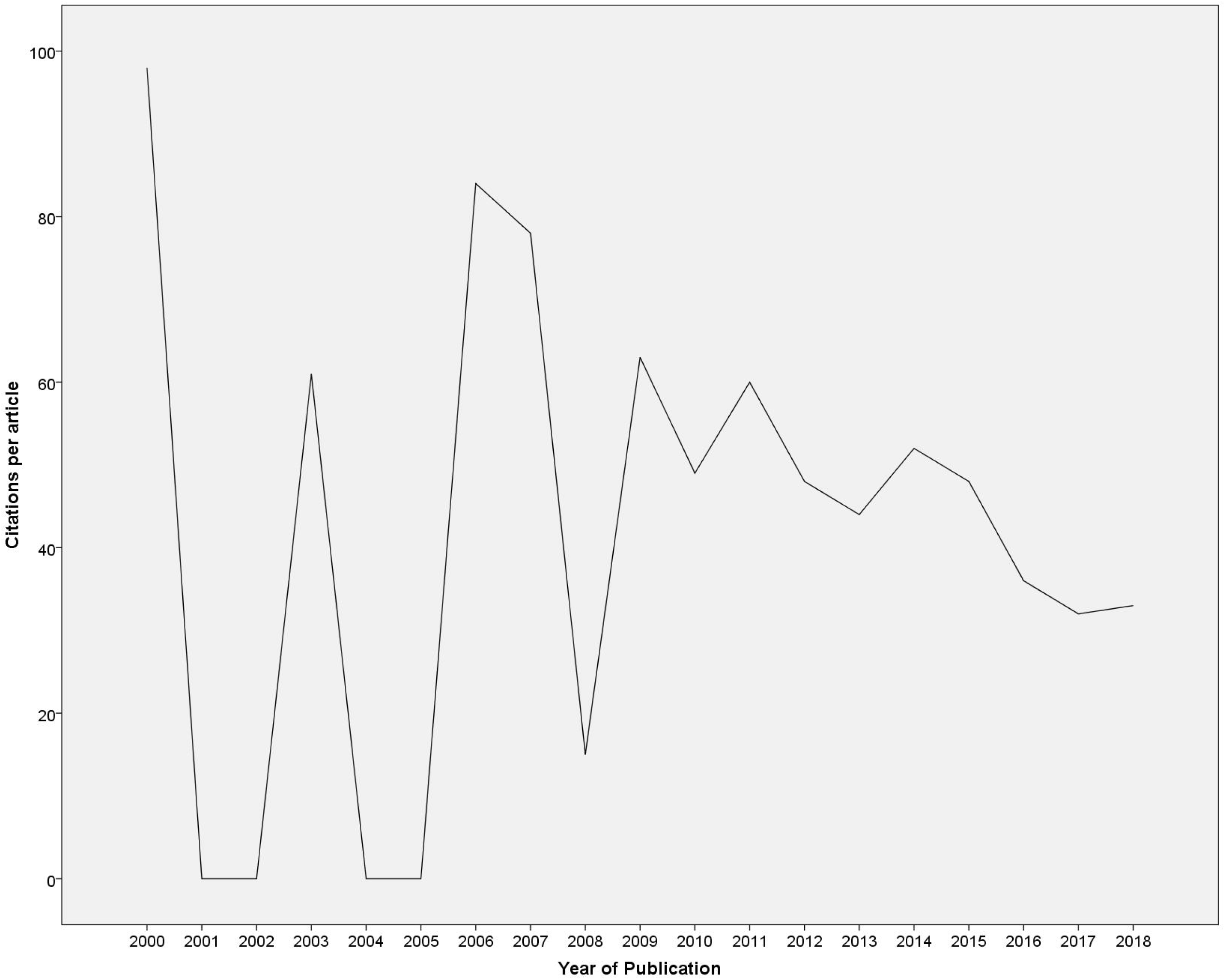
Number of citations per systematic review by year in OMFS.

**Figure 2.**
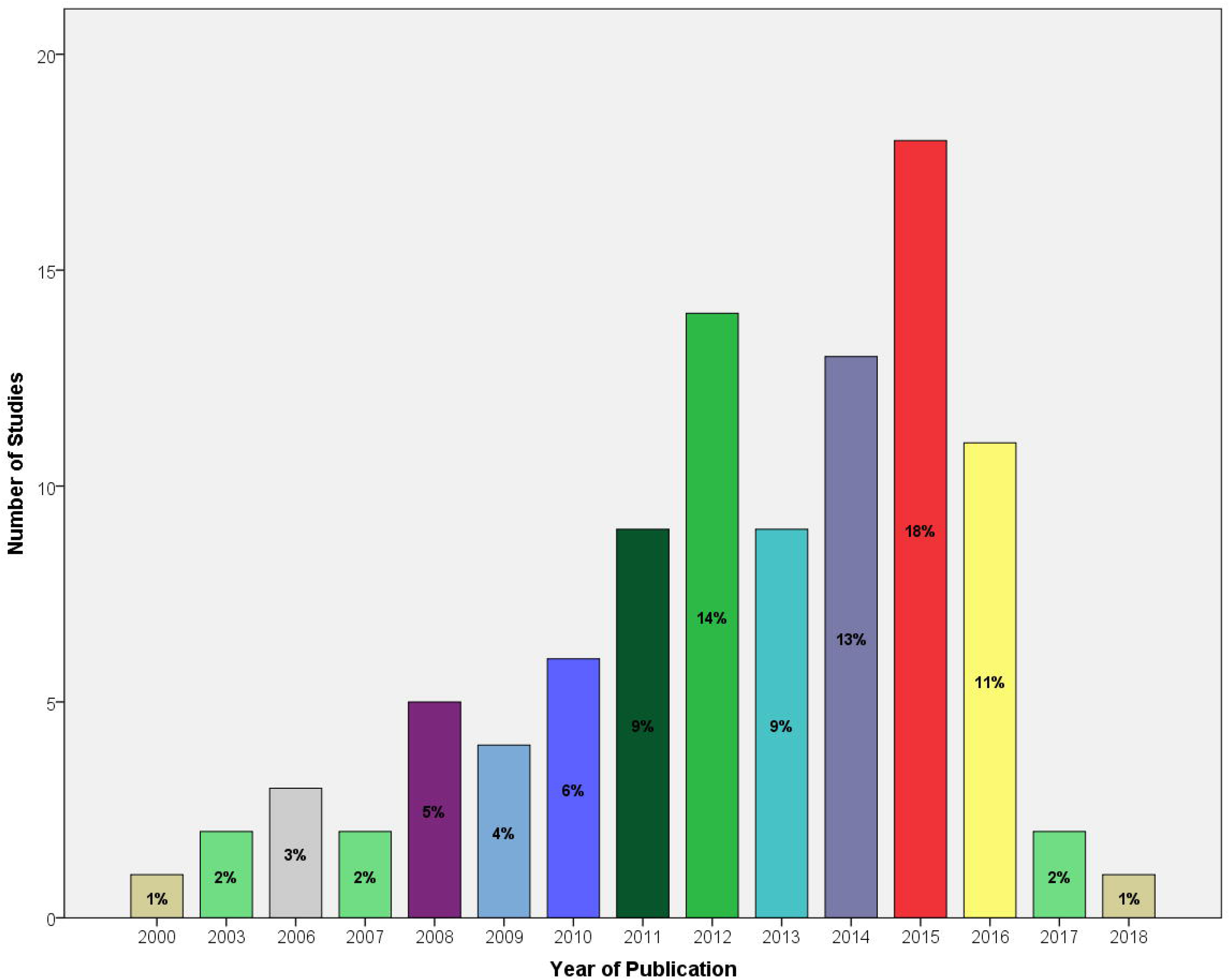
Time-pattern distribution of the 100 most cited systematic reviews on oral and maxillofacial surgery.

The topics “pre- and peri-implant surgery and dental implantology”, as well as “craniomaxillofacial deformities and cosmetic surgery” were the most frequently cited topics (22% each) in the top 1

00 list (**Figure 3**). The first topic has a total of 1334 citations while the second topic has a total of 1162 citations.

**Figure 3.**
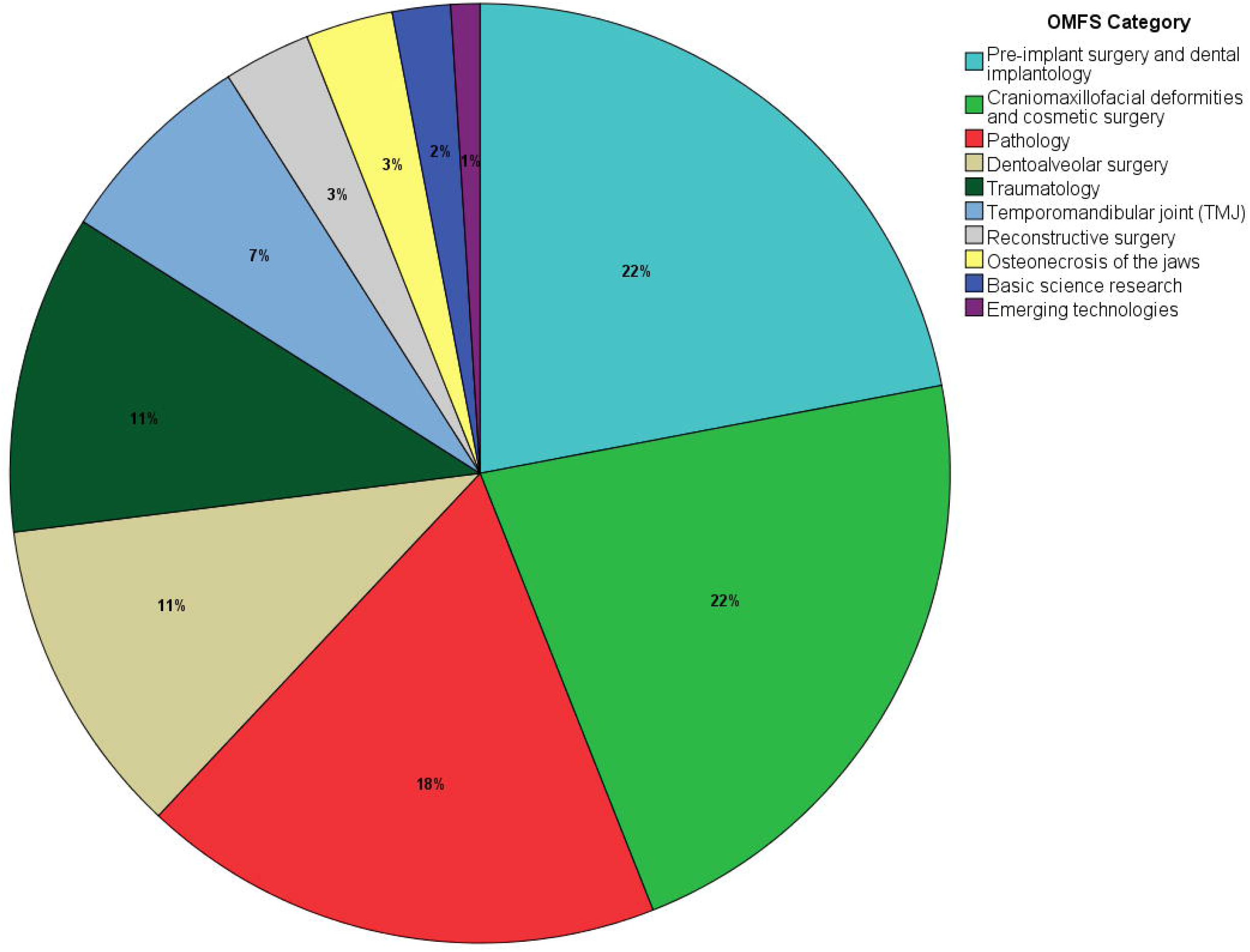
Topics covered among the 100 most cited systematic reviews on oral and maxillofacial surgery.

There were 25 different countries of origin and 83 institutions responsible for the highly cited systematic reviews. The leading countries were The Netherlands and Italy with 12 manuscripts each, followed by the USA with 10 articles (**Figure 4**).

**Figure 4.**
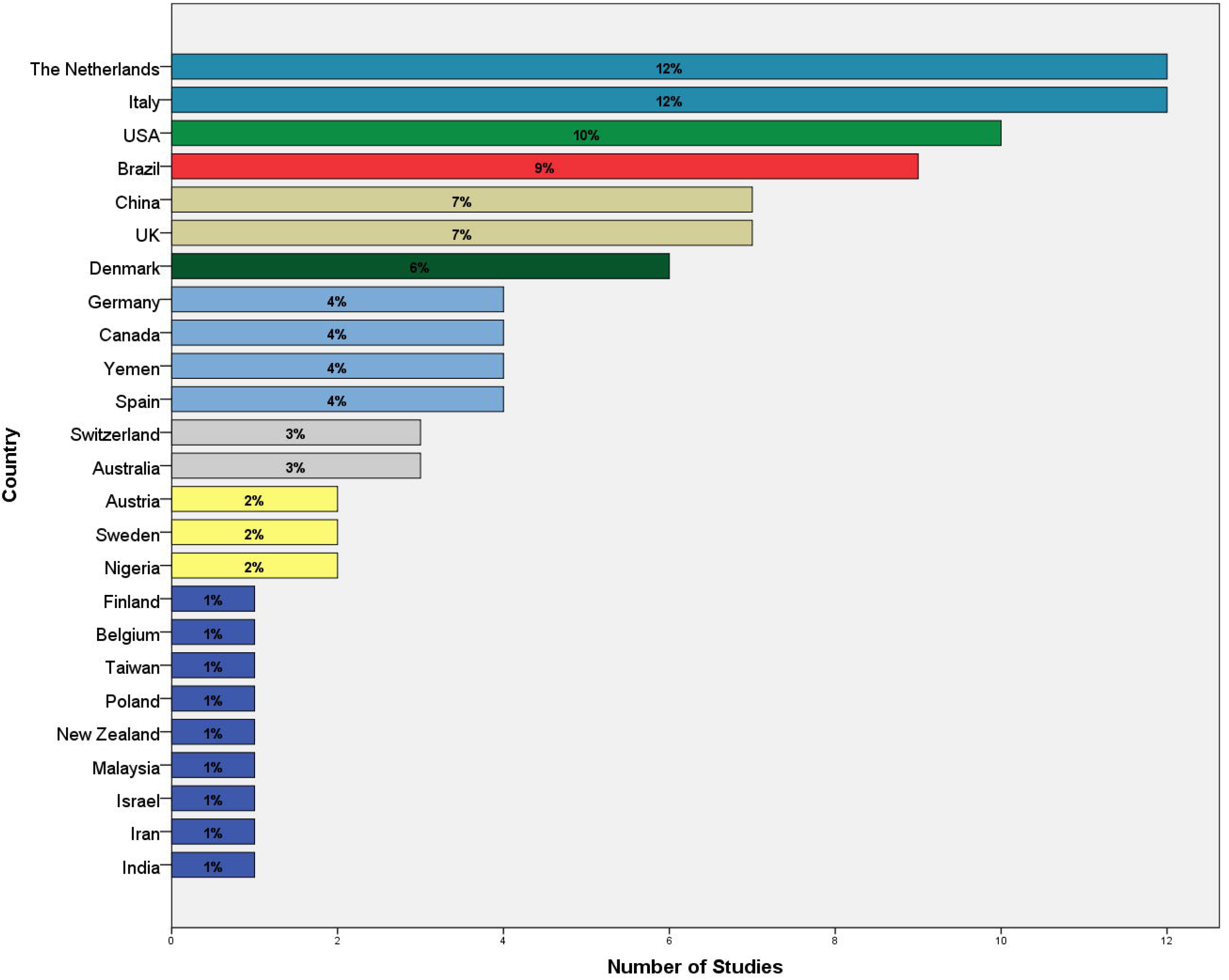
Country of author’s institute for 100 most cited systematic reviews.

Overall, IJOMS was responsible for 2481 citations, JOMS for 1684, TRIPLEO for 380 citations, JCMFS for 307, and BJOMS for 255. The mean citation rate per published review followed the same pattern with IJOMS having a mean citation rate of 57.7 citations per review, JOMS 49.5 citations per review, TRIPLEO 47.5 citations per review, JCMFS 38.4 citations per review, and BJOMS 31.9 citations per review. Most of the manuscripts were published in the IJOMS and JOMS (Figure 5).

**Figure 5.**
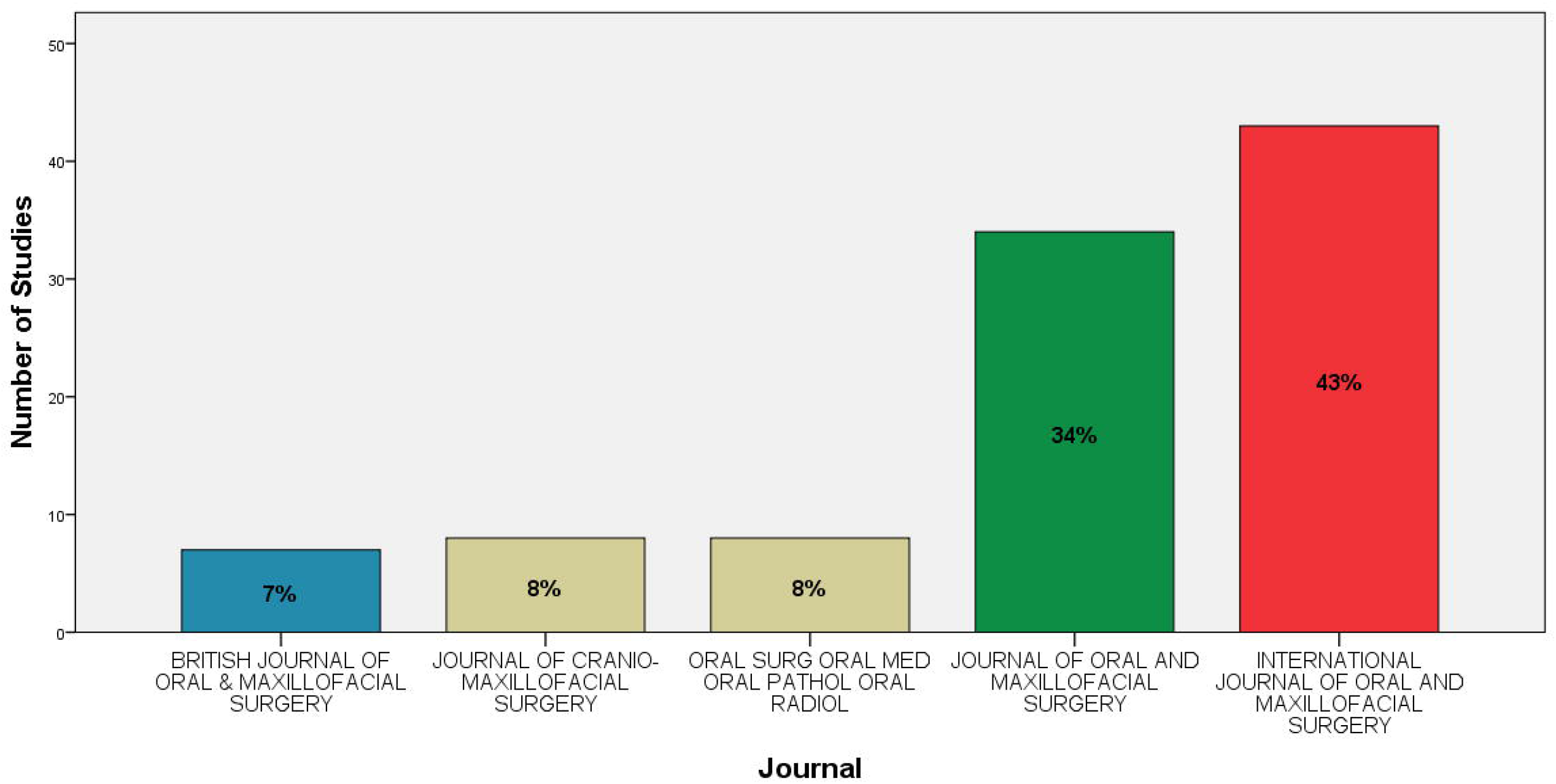
Most frequently cited journal.

## DISCUSSION

In the field of (dental)medicine, as in the other fields of science, there is wish reach out, to affect decisions, and to guide the reader in a decision making ^8^. Articles reaching more than 100 citations are considered classic, i.e. having a great impact ^12^. However, to analyze and understand if the conducted research has any impact or affects decision making one has to analyze how and if the articles do reach out, and which impact they have in their research filed ^12^. To do that scientometrics, which is bibliometry in the field of science, is frequently used. This study used citation analysis ^13^, to identify the publications that have had the greatest impact in the field of OMFS. Although the field of OMFS is very wide, with diverse conditions and treatments, the main finding of this citation analysis indicates that there were only 7 systematic reviews that reached to 100 citations in the field of OMFS, among the included journals. This is in line with previous studies indicating that less 10% of the published articles reaches up to the status of classic articles ^14-16^.

Among the classic articles the top cited article was an Italian systematic review on longitudinal studies about the evaluation of survival and success rates of dental implants ^9^, published in 2015 but already up in 200 citations. Among top-three there was one more systematic review on dental implants also from Italy focusing on different alveolar bone augmentation procedures for implant placement ^17^, published in 2014 and has now reached 124 citations. It is not surprising that systematic reviews upon dental treatments are top ranked since dental implant surgery is the vast most common surgical procedure next to tooth extractions, in contrast to orthognathic surgery, tumors.^18^

The second most cited was also a European (from The Netherlands) systematic review on three-dimensional image fusion processes for planning and evaluating orthodontics and orthognathic surgery, published in 2011 having 131 citations. All top-three systematic reviews were published in IJOMS, which is the top journal in the field of OMFS, being ranked in the first quartile (QR1). It has been shown that the journal impact factor answers for 59% of the variation in the number of citations ^19^. Therefore, it is not at surprising finding since most authors are interested in publishing papers in journals with high impact-factors, which also is considered an indication of high quality papers ^20,21^. Just outside top-three systematic reviews this citation analysis could show that systematic reviews upon osteoradionecrosis from Hong Kong (112 citations)^22^ and Malaysia (100 citations)^23^, as well as osteonecrosis from Germany (104 citations)^24^. The same topics were also dominating the rest of the most highly cited systematic reviews.

It has previously been reported that the majority of the top ranked, top cited publications are produced in nations with better economic rankings ^16,25^. This was also found in this citation analysis indicating that most of the systematic reviews in the field of OMFS are produced in Europe and the US, as well as Hong Kong, with Italy as the most successful country.

One interesting factor is that the top ranked systematic reviews are all published after the year 2010, however not surprising since only 14 out of the 100 top cited were published in the decade 2000-2010. One common criticism on citation analysis reports is that the outcome is affected by the impact of time ^14^. This was, however, not the case in this report on the field of OMFS. In accordance with our results, previous studies have indicated that there are just a few citations the first years, with a peak of citations just before an article-age of 10 years ^26^.

Another aspect to consider is that only can be used to assess the impact the specific article has on its field by quantifying its recognition, the importance, and also perhaps how common or severe a condition might be, but it cannot reflect the quality of the content in the article ^20,21^. Therefore, it is of great importance to use the outcome of this article as a source of information not just for researchers but also for clinicians and students, and which areas have a large impact on the field of OFMS.

## Data Availability

not required

